# Continued need for non-pharmaceutical interventions after COVID-19 vaccination in long-term-care facilities

**DOI:** 10.1101/2021.01.06.21249339

**Authors:** Jay Love, Lindsay T. Keegan, Frederick J. Angulo, John McLaughlin, Kimberly M. Shea, David L. Swerdlow, Matthew H. Samore, Damon J.A. Toth

## Abstract

Long-term care facilities (LTCFs) bear disproportionate burden of COVID-19 and are prioritized for vaccine deployment. LTCF outbreaks could continue occurring during vaccine rollout due to incomplete population coverage, and the effect of vaccines on viral transmission are currently unknown. Declining adherence to non-pharmaceutical interventions (NPIs) against within-facility transmission could therefore limit the effectiveness of vaccination. We built a stochastic model to simulate outbreaks in LTCF populations with differing vaccination coverage and NPI adherence to evaluate their interacting effects. Vaccination combined with strong NPI adherence produced the least morbidity and mortality. Healthcare worker vaccination improved outcomes in unvaccinated LTCF residents but was less impactful with declining NPI adherence. To prevent further illness and deaths, there is a continued need for NPIs in LTCFs during vaccine rollout.

## MAIN

The COVID-19 pandemic has been particularly devastating for residents of LTCFs (skilled nursing homes and assisted living facilities). While there has been a shift of disease burden to younger populations since the early months of the pandemic, deaths of people over 65 years of age and those in LTCFs continue to constitute large proportions of total COVID-19 deaths in the US (*1*). In advance of potential vaccine availability, discussion on the optimal vaccine allocation strategy has resulted in rollout plans to target priority populations. In the US, front-line healthcare workers and LTCF residents have been prioritized to receive the first-available vaccines (*2*). However, questions remain about the path by which vaccines may act to reduce COVID-19 spread and achieve herd immunity (*3*–*5*). These questions are especially relevant when considering an initially limited vaccine supply.

A critical factor that may complicate efforts to contain COVID-19 through vaccination is “pandemic fatigue”. This phenomenon, characterized by demotivation to follow recommended protective non-pharmaceutical interventions (NPIs) such as mask wearing and social distancing, has been implicated as a factor in recent surges in infectious spread (*6*). A significant concern is that the availability of a vaccine could lead to a perception among the public that the pandemic has ended, resulting in additional behavioral changes that undermine any potential vaccine-derived transmission reduction. Here, we highlight how both standing variation in NPI adherence and changes in NPI adherence over time can substantially alter the population-level effect of the vaccine on morbidity and mortality in LTCFs.

We developed a dynamic-network, agent-based model under a modified *Susceptible-Exposed-Infectious-Recovered* (SEIR) paradigm to simulate disease spread in LTCFs. To quantify vaccine effects, we calibrated our assumptions to represent 95% efficacy in preventing COVID-19 disease as achieved in vaccine clinical trials (*7*–*9*). We considered the possibility that the vaccine is less than 95% effective in preventing SARS-CoV-2 infection, which could be consistent with clinical trial results (*9*) if vaccinated individuals had a higher rate of asymptomatic infection. Specifically, our model explicitly incorporates two pathways by which a vaccine may influence disease dynamics: 1) by reducing susceptibility to infection (by factor Ψ) and 2) by reducing disease progression (e.g., reducing symptoms by factor μ). These values were constrained to satisfy the overall vaccine effectiveness equation 1 – (1 – Ψ)(1 – μ) = 0.95. We conducted simulations with random pairs of the two vaccine effectiveness components and random levels of vaccine coverage in healthcare worker and resident populations. We simulated three scenarios for levels of NPI adherence, quantified by possible facility-specific rates of transmission-relevant resident-to-resident contact. Scenario 1 reflected strict NPI adherence among residents (physical distancing, mask wearing, restricted social mixing, etc.), assuming average resident-to-resident contact rate of 2 contacts per day per resident. Scenario 2 reflected gradual reduction in NPI adherence over time. Scenario 3 reflected low NPI adherence, with unrestricted resident-to-resident mixing quantified as 50 contacts per day per resident. An additional baseline scenario simulated no vaccine intervention but strict adherence to social distancing guidelines.

In the model, populations of residents and healthcare workers approximated US national averages (*10*), with 100 residents and 51 healthcare workers in each simulation. Simulations started with fully susceptible facility populations, and all importations of SARS-CoV-2 came through community contacts of healthcare workers and residents. After a non-infectious latent period, infectious periods were either fully asymptomatic or progressed through a pre-symptomatic state before symptom onset. Pre-symptomatic and asymptomatic infectious individuals could transmit with the same average infectiousness assumed for both states. Individual transmission rates were randomized by assigning each individual an infectiousness factor drawn from a gamma distribution to approximate individual variation in viral shedding (*11*). Symptomatic healthcare workers were immediately furloughed upon symptom onset, preventing subsequent transmissions in the facility, while symptomatic residents could continue transmitting to facility contacts at a 90% reduced rate, assuming additional precautions. Symptomatic infections were partitioned into mild and severe infections, and a portion of severe infections resulted in hospitalization or death, at frequencies commensurate with data for LTCF residents and healthcare workers. Each simulation ran for 100 days with discrete daily time steps.

We found that all disease outcomes (number of infections, severe infections, hospitalizations, and deaths) were highest in the absence of a vaccine (the baseline scenario) and lowest within LTCFs when vaccine deployment was paired with strong adherence to NPIs (Scenario 1; 52% fewer infections and 67% fewer severe infections, hospitalizations, and deaths than baseline; Figure 1). Levels of morbidity and mortality for scenarios 2 and 3 (declining NPI adherence and stable but low NPI adherence, respectively) were intermediate between those of scenarios 1 and baseline (scenario 2: 17% fewer infections, 43% fewer severe infections and hospitalizations, and 44% fewer deaths than baseline; scenario 3: 15% fewer infections and 41% fewer severe infections, hospitalizations, and deaths than baseline). Notably, simulations with a gradual reduction in NPI adherence (Scenario 2) yielded results that were more similar to simulations with low NPI adherence (Scenario 3) than to those with high NPI adherence (Scenario 1), indicating that pandemic fatigue could significantly undermine the impact, in terms of morbidity and mortality, of a vaccine rollout if not anticipated and accounted for.

**Figure 1.**
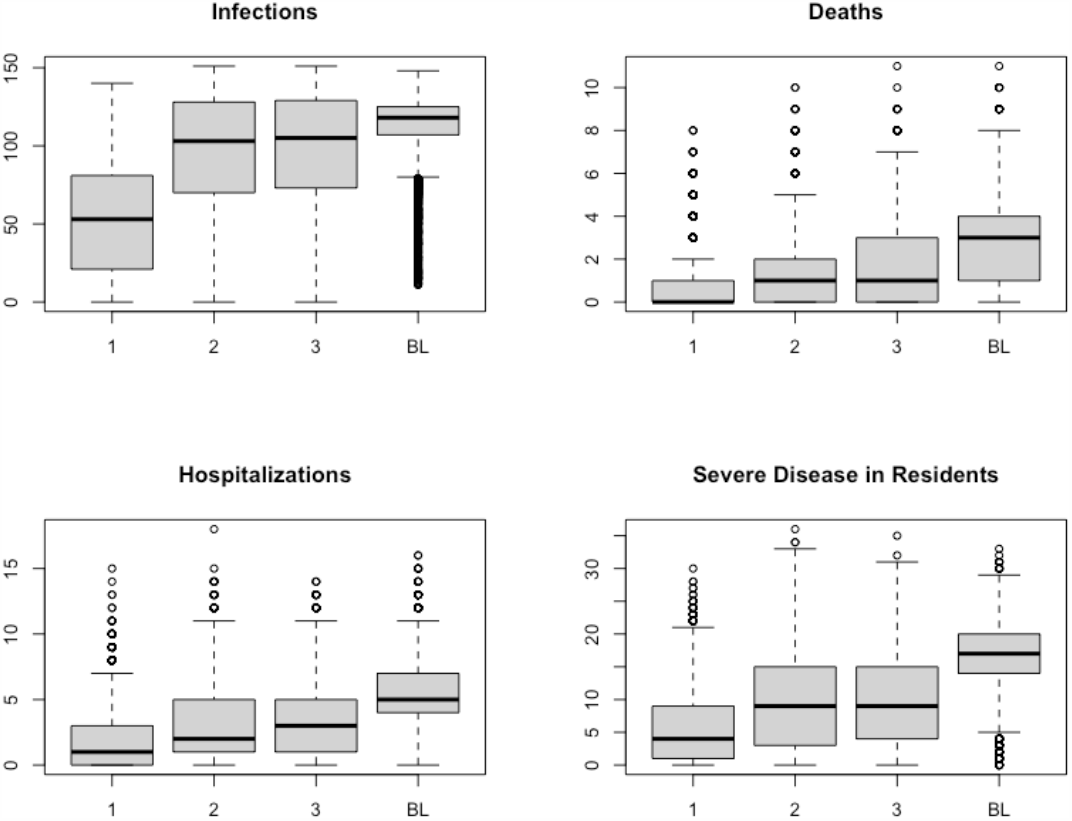
Disease outcomes by scenario. Comparison of total infections, severe infections in residents, hospitalizations, and deaths over 100 days in simulated populations of 100 residents and 51 healthcare workers at long-term care facilities. Scenarios 1, 2, and 3 paired vaccination with strong, gradually reduced, and weak adherence to NPIs, respectively. Scenario BL (baseline) included no vaccination but strong adherence to NPIs. Boxplots show median, 1^st^ and 3^rd^ quartile, and whiskers extend to 1.5 times the interquartile range. Statistical outliers included as open circles.

The impact of increasing vaccine coverage among healthcare workers compared to residents in LTCFs differed across NPI adherence scenarios. In all scenarios, increasing vaccine coverage of both healthcare workers and residents prevented morbidity and mortality, but the prevention was largest when NPI adherence was high (Scenario 1). Moreover, degree of NPI adherence strongly modulated the impact of increasing healthcare worker vaccination coverage on prevention of deaths. By contrast, the impact of increasing resident vaccination coverage on prevention of deaths was more robust to diminished NPI adherence. As is shown in Figure 2, increasing healthcare worker vaccination coverage (horizontal axes, Figure 2) prevents deaths, as does increasing resident vaccination coverage (vertical axes, Figure 2). However, the per-vaccine rate of death prevention attributable to healthcare worker vaccination was markedly lower for scenarios without high NPI adherence. The per-vaccine rate of death prevention attributable to resident vaccination was also lower without strong NPI adherence, but this prevention was less pronounced than that observed in reference to health-care worker vaccination. This finding indicates that, in LTCFs with weak NPI adherence, strategies that prioritize vaccinating healthcare workers may have only a weak effect on prevention of COVID-19 disease burden in LTCFs. In comparison, strategies that prioritize resident vaccination are more stable to differing adherence to NPIs.

**Figure 2.**
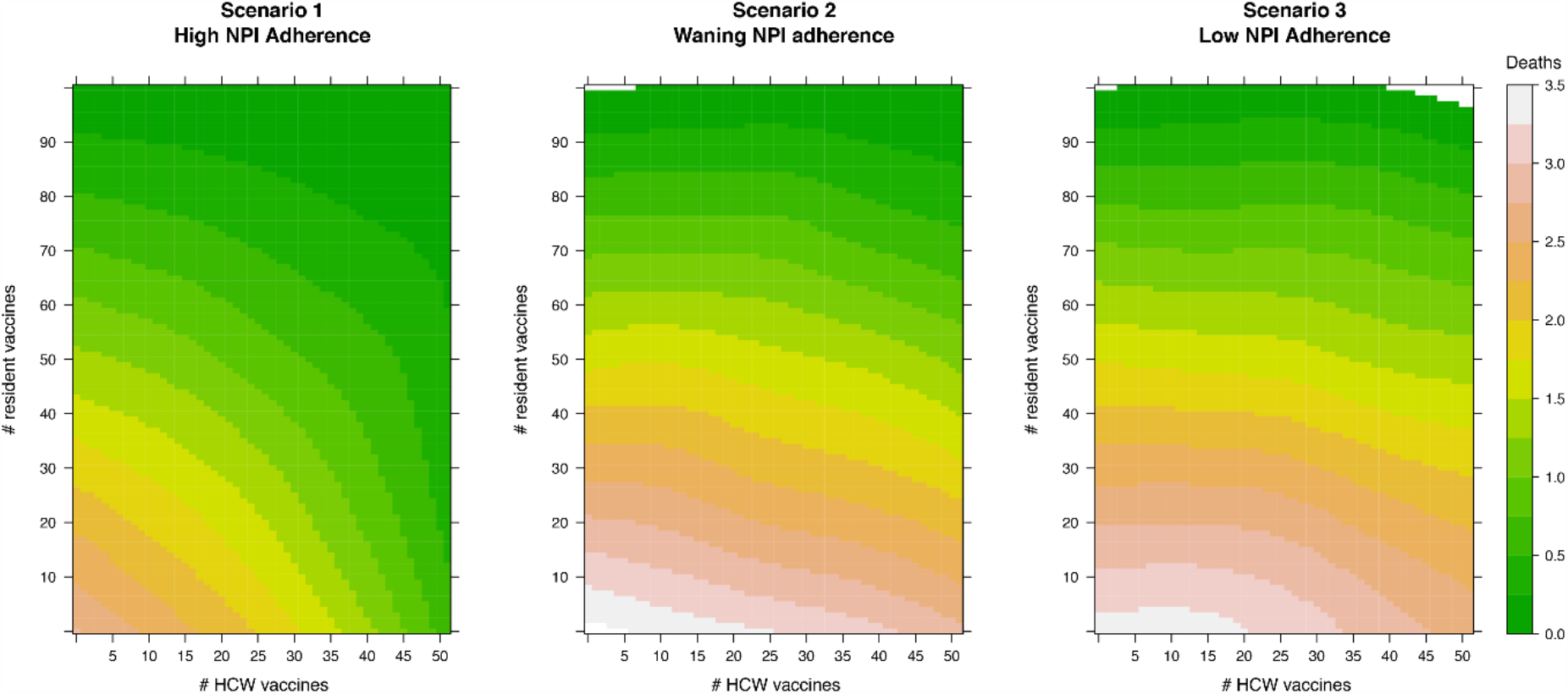
Impact of vaccine coverage on COVID-19 deaths. Heatmaps of the impact of vaccine coverage on COVID-19 deaths in simulated long-term care facility populations of 51 healthcare workers and 100 residents for three scenarios with different NPI adherence. Warmer colors indicate more deaths. Per vaccine, vaccinating healthcare workers prevents more deaths when NPI adherence is high (Scenario 1, left), but that impact declines substantially when NPI adherence wanes (Scenario 2, middle) or is low (Scenario 3, right).

It is important to address why the impact of increasing healthcare worker vaccination coverage would be strongly dependent upon NPI adherence while the impact of increasing resident vaccination coverage would be less so. Vaccine efficacy measured in clinical trials quantifies the reduction of symptomatic disease in the vaccinated group versus the placebo group (*9*), and it is currently unknown the relative extent to which that efficacy is derived from a reduction in susceptibility to infection versus a reduction in disease progression after infection has occurred. We designed our model to account for all possible ways that vaccine-induced reduction in symptomatic disease could arise. It is possible that vaccine efficacy is strongly derived from a reduction in susceptibility, and our model incorporates that possibility. However, if reported vaccine efficacy is derived partially through reduction in progression to symptomatic disease, vaccination could involve an inadvertent increase in the proportion of infections that are asymptomatic, as has been discussed in theoretical work referencing other diseases and their vaccines (*5, 12*). It is in simulations where the vaccine both blocks infection and reduces progression to symptomatic disease that, while vaccination results in a reduction in total infections, vaccinated, asymptomatic infections are still infectious, leading to additional burden among unvaccinated individuals who could develop symptomatic disease. If those unvaccinated individuals are more prone to severe disease outcomes, as are residents in LTCFs when compared to healthcare workers, then population-level morbidity and mortality may not experience the expected decline associated with vaccination. Simulations that are prone to this phenomenon are likely to be largely responsible for the observed pattern of dependency of the impact of healthcare worker vaccination coverage on NPI adherence.

For this reason and also because healthcare worker vaccination does not address community-to-resident infections (e.g., through visitation, sojourns, or facility transfer), vaccinating only healthcare workers and relying on indirect effects to reduce resident deaths may not be as impactful as strategies that prioritize resident vaccination. This potential challenge can be mitigated by continued strict adherence to NPIs that reduce resident-to-resident contact until high vaccine coverage is achieved in both populations. As more is learned about the relative contribution of the components of COVID-19 overall vaccine efficacy, such possibilities could prove to be only minor considerations. Until that time, the findings of our study indicate that care should be taken to carefully consider adherence to NPIs when determining initial allocation of COVID-19 vaccine in LTCFs.

Our model provides a useful tool to evaluate the allocation of a limited allotment of vaccine with punctuated deployment, especially in the context of reduced or relaxing NPI measures. Here we demonstrate that vaccinating LTCF residents will likely lead to meaningful reductions in morbidity and mortality. However, guidance to preferentially vaccinate healthcare workers, while suitable for many applications, may not be optimal for reducing COVID-19 morbidity and mortality in LTCFs that have difficulty adhering to NPIs. Our results suggest that maintaining adherence to NPIs is essential to reducing COVID-19 burden in LTCFs, especially until more is known about the impact of vaccines on transmission. Particularly in the face of pandemic fatigue, limiting high-risk interactions between residents and maintaining limitations on visitors is essential to support vaccination efforts. Additionally, individual facility allocation strategies should account for facility-specific adherence to NPIs. LTCFs that have reduced ability to limit resident-resident contact, such as those catering to dementia patients, should ensure that vaccine allocation prioritizes a reduction in the most important endpoint and supports those who can safely distance. Overall, there is a continued need for NPIs in LTCFs, including after COVID-19 vaccination has commenced.

## Data Availability

Data and model code will be made publicly available upon publication or request.

Drs. Angulo, Mclaughlin, Shea, and Swerdlow reported being employed by Pfizer Vaccines. This work was supported by Pfizer. Pfizer Inc. reviewed this manuscript and approved the decision to submit the manuscript for publication.

## References

1. J. A. W. Gold, L. M. Rossen, F. B. Ahmad, P. Sutton, Z. Li, P. P. Salvatore, J. P. Coyle, J. DeCuir, B. N. Baack, T. M. Durant, K. L. Dominguez, S. J. Henley, F. B. Annor, J. Fuld, D. L. Dee, A. Bhattarai, B. R. Jackson, Race, Ethnicity, and Age Trends in Persons Who Died from COVID-19 — United States, May–August 2020. MMWR Morb Mortal Wkly Rep. 69, 1517–1521 (2020).

2. K. Dooling, N. McClung, M. Chamberland, M. Marin, M. Wallace, B. P. Bell, G. M. Lee, H. K. Talbot, J. R. Romero, S. E. Oliver, The Advisory Committee on Immunization Practices’ Interim Recommendation for Allocating Initial Supplies of COVID-19 Vaccine — United States, 2020. MMWR Morb Mortal Wkly Rep. 69, 1782–1786 (2020).

3. S. H. Hodgson, K. Mansatta, G. Mallett, V. Harris, K. R. W. Emary, A. J. Pollard, What defines an efficacious COVID-19 vaccine? A review of the challenges assessing the clinical efficacy of vaccines against SARS-CoV-2. Lancet Infect Dis. 3099, 1–10 (2020).

4. B. M. Lipsitch, N. E. Dean, Understanding COVID-19 vaccine efficacy. Science (80-). 5938, 1–5 (2020).

5. S. Gandon, M. J. Mackinnon, S. Nee, A. F. Read, Imperfect vaccines and the evolution of pathogen virulence. Nature. 414, 751–756 (2001).

6. World Health Organization, Pandemic fatigue: Reinvigorating the public to prevent COVID-19 (Copenhagen, 2020).

7. P. Doshi, Covid-19 vaccine trial protocols released. BMJ. 371, m4058 (2020).

8. E. Mahase, Covid-19: What do we know about the late stage vaccine candidates? BMJ. 371, m4576 (2020).

9. F. P. Polack, S. J. Thomas, N. Kitchin, J. Absalon, A. Gurtman, S. Lockhart, J. L. Perez, G. P. Marc, E. D. Moreira, C. Zerbini, R. Bailey, K. A. Swanson, S. Roychoudhury, K. Koury, P. Li, W. V Kalina, D. Cooper, R. W. Frenck Jr., L. L. Hammitt, Ö. Türeci, H. Nell, A. Schaefer, S. Ünal, D. B. Tresnan, S. Mather, P. R. Dormitzer, U. Şahin, K. U. Jansen, W. C. Gruber, Safety and Efficacy of the BNT162b2 mRNA Covid-19 Vaccine. N Engl J Med, 1–13 (2020).

10. C. S. Gabriel, “An overview of nursing home facilities: Data from the 1997 National Nursing Home Survey” (Hyattsville, Maryland, 2000).

11. X. He, E. H. Y. Lau, P. Wu, X. Deng, J. Wang, X. Hao, Y. C. Lau, J. Y. Wong, Y. Guan, X. Tan, X. Mo, Y. Chen, B. Liao, W. Chen, F. Hu, Q. Zhang, M. Zhong, Y. Wu, L. Zhao, F. Zhang, B. J. Cowling, F. Li, G. M. Leung, Temporal dynamics in viral shedding and transmissibility of COVID-19. Nat Med. 26 (2020), doi:10.1038/s41591-020-0869-5.

12. S. Gandon, M. Mackinnon, S. Nee, A. Read, Imperfect vaccination: Some epidemiological and evolutionary consequences. Proc R Soc B Biol Sci. 270, 1129–1136 (2003).

