## supplement for "Continued need for non-pharmaceutical interventions after COVID-19 vaccination in long-term-care facilities"

**<u>Supplemental Materials</u>**

### Methods

We developed a dynamic-network, agent-based model under a modified *Susceptible-Exposed-Infectious-Recovered* (SEIR) paradigm to simulate disease spread in LTCFs. The model was built in the R statistical environment [1]. It uses a nested-loop structure which, while relatively computationally expensive, is highly intuitive and allows for individual-level record keeping and dynamics that scale up to stochastic population-level patterns. The model can be understood as two interacting components: the dynamic network model and the disease transmission model.

#### *The dynamic network model*

Upon initiation of a model simulation, three undirected contact matrices are generated: one for resident-resident contacts, one for worker-worker contacts, and one for resident-worker contacts. Together, these matrices describe a network of 100 residents and 51 workers. The initial condition for the resident network contains randomly generated contacts with mean degree of 2. The worker network is subdivided into 3 “shifts” to simulate more realistic coworker contact. Contacts in the worker network are also randomly generated, but contacts are exclusively between workers of the same shift. Probability of contact with a non-shiftmate is 0, whereas probability of contact with a shiftmate is 0.47, resulting in a mean degree of 8 in the worker network. Resident-worker contacts in the interaction matrix are generated randomly with probability based on the designated mean degree and node number of resident and worker matrices.

The network changes over the course of a simulation. Each timestep (day) begins with regeneration of each network, updating contacts randomly following the rules outlined above. No cohorting is assumed and the network is memory-less (i.e., past states do not influence future states). Workers may be removed from the network due to quarantine away from the facility (as described below in the disease transmission model section) by zeroing their contacts for that day, and their duties are assumed by one-day, non-repeating, non-infectious substitutes drawn from outside of the existing population of workers in the network. In this way, quarantined workers cannot spread disease but their absence does not impact features of the network other than decreasing the connectivity of would-be contacts of quarantined workers. Quarantining of residents is handled in a different manner (see below).

Residents and workers have contacts outside of the facility. Each resident has the same fixed daily probability of visitation (1/7, or an average of one outside contact per week). Worker degree of outside contact is heterogenous; each worker is assigned a level of outside contact drawn from a uniform distribution between 0 and 4.

*The disease transmission model*

Individuals can move between disease states (*Susceptible-Exposed-Infectious-Recovered*), which are explicitly designated at the individual level within the model, according to model parameters (Table S1). While disease transmission in the model occurs through daily application of an algorithm iterated across individuals rather than through a system of population-level mathematical equations, as would be in a traditional SEIR model, the model that we use can be reasonably visualized in the diagram in Figure S1. A key feature of the model designed to reflect COVID-19 disease dynamics is the split between the exposed-to-asymptomatic infectious and the exposed-to-presymptomatic infectious paths. The baseline probability of becoming presymptomatic was 60%, following CDC modeling recommendations.

Individuals were quarantined if they developed symptomatic disease. Each symptomatic infection was assigned a score between 0 and 20 for disease severity. Since younger people tend to have lower rates of severe disease than older people, and since workers tend to be much younger than residents, worker disease severity for symptomatic cases was drawn from a normal distribution centered at 9 with a standard deviation of 2, while resident disease severity was drawn from a normal distribution centered at 15 with a standard deviation of 2. A cutoff value was established for the designation of severe disease at a severity score of 15. In this way, about half of symptomatic cases in residents become severe, while workers only rarely develop severe disease. Hospitalization cutoff value of severity was 17. Different death rates were defined for workers and residents and a daily death rate was derived considering length of infection for severe cases (see Table S1).

Individuals are quarantined upon development of symptoms. In contrast to quarantine of workers (as described above), quarantine of residents is "leaky". For any contacts involving a symptomatic infectious resident, the infection rate parameter was reduced by a factor of 0.9.

Starting conditions assumed a completely naive population. Infectious spread begins when an individual is infected by a contact outside of the facility. Levels of outside contact described in the previous section determined the outside contact rate for each individual. Probability of an infection occurring though outside contacts was determined each day for each individual as a factor of the baseline infection rate, the level of outside contact for that individual, and the community prevalence of infectious COVID-19 (a value of 0.018 for these simulations, which reflects community prevalence in Salt Lake City, UT in early December, 2020).

One of our goals was to evaluate the impact of a vaccine with 95% efficacy as defined in clinical trials [2]. This level of efficacy to prevent symptomatic disease could arise through a combination of prevention of infection and prevention of progression to symptomatic disease. We implemented these two potential modes of vaccine effectiveness in the model as distinct parameters that relate to each other through their joint contribution to vaccine efficacy: [$1 - (1 - \psi)(1 - \mu) = 0.95$]. These factors modified probability of infection and probability of symptom development for vaccinated individuals, respectively. Residents had a 5% reduction in the effectiveness of these parameters. Vaccination coverage levels were determined at the start of each simulation and did not change across the duration of the simulated epidemic.

*Visualizations*

Figure 2 depicts heatmaps that compare effects of vaccine coverage levels on deaths across three scenarios. These heatmaps were created by fitting a second-degree loess smoothing function with a span value of 0.5 to the data and interpolating to all values within the data range, then producing plots with the levelplot function from the lattice package in R [3].

**References**


1. R Core Team. R: A language and environment for statistical computing. (2017).
2. Polack, F. P. *et al.* Safety and Efficacy of the BNT162b2 mRNA Covid-19 Vaccine. *N Engl J Med* 1–13 (2020). doi:10.1056/NEJMoa2034577
3. Sarkar, D. *Lattice: Multivariate Data Visualization with R*. (Springer, 2008).

Table S1. Parameter values.

| parameter | value | notes |
|---|---|---|
| n residents | 100 | |
| n workers | 51 | health care workers were split into three shifts per day, members were randomly selected |
| n resident-resident contacts per resident per day | 2 | randomly generated contacts each day |
| n worker-worker contacts per worker per day | 8 | random contacts drawn from shiftmates each day |
| resident-worker contacts | *see note | resident-worker contacts generated from a probability of relative populations with the criteria that each resident was contacted by >=1 worker per day |
| infectiousness | 0.11 | per-contact rate of infection |
| duration of infection | 7 | days |
| duration of severe infection | 14 | days |
| infection latency | 3 | days |
| proportion of infections that become symptomatic | 0.6 | |
| healthcare worker death rate | 0.0002 | for symptomatic cases |
| resident death rate | 0.25 | for symptomatic cases |
| community prevalence | 0.018 | based on levels of community prevalence in Salt Lake City in early December, 2020 |
| resident visitation rate | 0.14 | daily probability of community contact for each resident |
| worker visit rate | *see note | number of daily contacts for each resident was stable across an epidemic simulation. Values were drawn from a uniform distribution between 0 and 4. |
| vaccine efficacy | 0.95 | |
| vaccine efficacy reduction factor in elderly | 0.95 | applied to vaccine effects in residents |

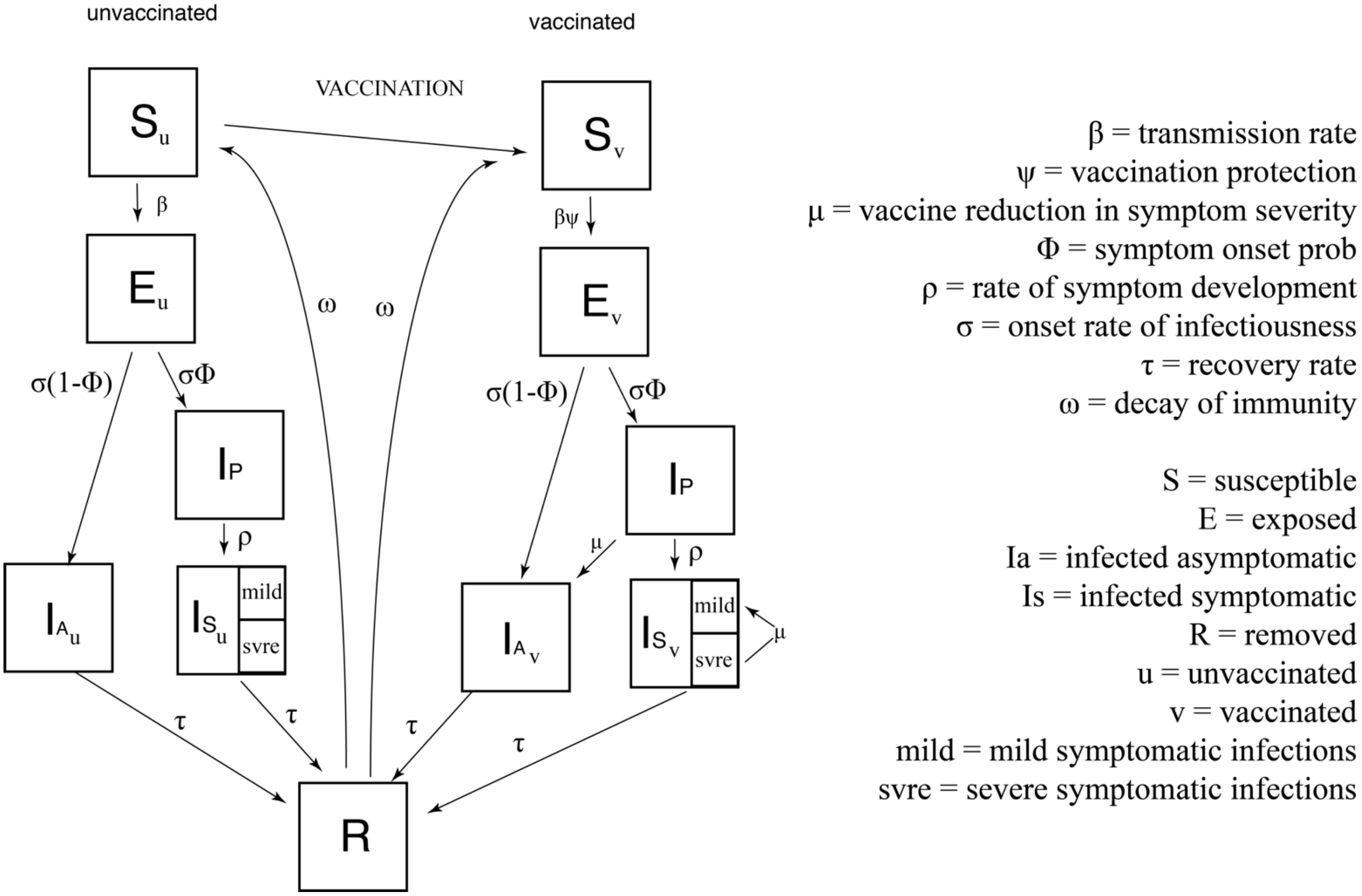


Figure S1. Model diagram. Individuals move between S, E, Ia, Ip, Is, and R compartments according to the rate parameters, applied as daily probabilities at each time step. Vaccination can affect dynamics of the system through $\psi$ and $\mu$.
